# Access to hypertension services and health-seeking experiences in rural Coastal Kenya: A qualitative study

**DOI:** 10.1101/2025.02.10.25321754

**Authors:** Robinson Oyando, Nancy Kagwanja, Brahima A. Diallo, Syreen Hassan, Jainaba Badjie, Ruth Lucinde, Noni Mumba, Samson Muchina Kinyanjui, Pablo Perel, Anthony Etyang, Edwine Barasa, Ellen Nolte, Benjamin Tsofa, IHCoR-Africa Collaborators

## Abstract

Globally, hypertension causes 10.8 million deaths annually. However, in Kenya, like in other low-and middle-income countries, access to hypertension care remains limited and inequitable. Understanding patients’ journeys in accessing care along the care cascade is critical to inform patient-centred care and policy improvements. This evidence is limited in Kenya. This study aimed to explore patient journeys in accessing hypertension care in rural Coastal Kenya―a setting with a high hypertension burden. We conducted a qualitative cross-sectional study and collected data using in-depth interviews (n=24) and focus group discussions (n=5) with hypertension patients and their adult family caregivers in two purposively selected sub-counties in Kilifi County. We conducted and transcribed the interviews in Kiswahili and Giriama (local languages) and translated them into English. We used QSR NVivo 12 for data management. We analysed the data using a framework approach and interpreted our findings using Levesque’s access framework. Access to hypertension screening and diagnosis services was undermined by information barriers which led to inadequate awareness and lack of knowledge about hypertension and its causes. There were perceptions of inadequate health facility capacity to offer hypertension screening services, particularly to ‘healthy’ individuals thus presenting as a barrier to demand for screening services. Acceptability of care was undermined by inadequate patient counselling at diagnosis and perceived disrespectful treatment of patients. Access to treatment and diagnostic tests was undermined by unaffordable care, limited availability of medicines and equipment, long waiting times, and inaccessible health facilities. Having health insurance enabled access to care, but most participants did not have a cover. Participants adapted to these access barriers by reducing/skipping daily medication doses, resorting to alternative forms of care (e.g., herbal treatments and faith healing), and changing health facilities for routine clinic appointments. Access to care for older patients and those with complications was enabled by family caregivers who coordinated and navigated the health system on their behalf. People living with hypertension experience a combination of interacting individual, community, and health system-related barriers to accessing care. There is a need to systematically address identified barriers and ensure patient-centred responses that meet patients’ needs. Strengthening the health system’s capacity to ensure availability and affordability of treatment and diagnosis services, creation of community hypertension awareness, adequate patient counselling at screening and diagnosis, and involvement of family caregivers for elderly patients are examples of urgent interventions to improve access to hypertension care.

## 1. Background

Hypertension is a major modifiable risk factor for cardiovascular diseases and the leading cause of preventable morbidity and mortality (1); in 2019, hypertension was implicated in one in five deaths and 9.3% of disability-adjusted life-years lost globally (2–4). Worldwide, the prevalence of hypertension in adults doubled from 650 million in 1990 to 1.3 billion in 2019 (5). Low- and middle-income countries (LMICs), including countries in sub-Saharan Africa (SSA), bear a disproportionate burden of hypertension. In 2019, approximately 700,000 deaths in SSA were attributed to hypertension, double the number seen in 1990 (2). It is estimated that 36% of adults in SSA who are aged between 30-79 years are hypertensive (1).

The rising burden of hypertension is a concern in LMICs as it widens existing inequalities, burdens households economically, and places further pressure on already resource-constrained and weak health systems (6–8). SSA, in particular, lacks the health system capacity to effectively detect and manage hypertension, with for example, Geldsetzer *et al.* finding that among individuals with hypertension, only 52% had ever had their blood pressure measured, 22% were diagnosed, 13% were on treatment, and 4.3% attained blood pressure control (9). Hypertension is a particular concern in rural SSA, where more than half of the population resides, and levels of awareness, treatment, and control are lower compared to urban settings (10–12).

Tackling the increasing burden of hypertension is a global health imperative. However, evidence suggests that the global target to reduce hypertension prevalence by 25% by 2025 is unlikely to be attained in many countries (13). Kenya, like other countries in SSA, has a high and increasing prevalence of hypertension. The estimated prevalence of hypertension among adults in Kenya is 33% (14), with the majority (73.1%) of people living with hypertension not on treatment and nearly half (48%) of those on treatment not attaining blood pressure control ((15). The implications of increasing hypertension burden are most felt in rural and poor settings, where access to needed medical care is limited and inequitable (16–22).

There is growing evidence of the complex and multiple access barriers to the effective management of major non-communicable diseases (NCDs), such as hypertension. For brevity, these can be categorised as micro (patient/community), meso (provider/health system), and macro (national context) barriers. At the micro level, key issues are the asymptomatic nature of hypertension, a lack of understanding of long-term risk, misconceptions about the benefits of pharmacotherapy, opportunity costs associated with seeking treatment, lack of adherence, and inadequate capacity to afford care (16, 18, 23–26). Meso-level barriers include inadequate systematic screening and early detection programs, poor provider communication (within and across different levels of care) and between providers and patients, lack of skills and competencies, poor infrastructure, erratic supply of antihypertensive medicines, and lack of adequate referral systems (27–31). Macro-level barriers include poor access to healthcare facilities, inadequate prepayment financing arrangements, underinvestment in health service capacity, and the apparent under-prioritization of NCDs in government budgetary allocations (32–37).

Effective and patient-centred responses to hypertension care require an understanding of patient and health system-related factors acting at each stage of the patient journey―from awareness, screening, diagnosis, treatment, follow-up, and control (26, 30, 38, 39). Understanding these factors from the patient’s perspective could help not only in improving access to care, but also in providing care models that are responsive to patients’ unmet needs, improve their participation in their management, and attain treatment targets (38, 39). Such context-specific evidence is limited in SSA, including Kenya (with notable exceptions, such as the study by Naanyu *et al.* which explored patient experiences with an integrated service model for people with chronic conditions in Western Kenya) (40). The study found that individuals’ financial resources, their social situation at each stage of the journey and specific system context significantly affected the patient journey (40). However, there is a need for further research that looks at experiences in other contexts and settings. Moreover, the empirical literature has explored experiences more broadly, and in some instances, focused on one stage in the patient journey, often treatment (31, 41). Finally, the extant literature on access barriers experienced by NCD patients has primarily been conducted in high-income settings, with few studies in rural settings in SSA (20, 42). This study seeks to contribute to closing this evidence gap by systematically examining the experiences of people with hypertension in accessing primary care services along the care cascade. We conceptualised access in broad dimensions to integrate demand and supply factors of access in a rural setting with a high hypertension burden in Coastal Kenya.

## 2. Methods

This study is part of a broader project: Improving Hypertension Control in Rural Sub-Sharan Africa (IHCoR-Africa) that aims to co-develop and evaluate a community-centred approach to improve the management of hypertension for people living with hypertension in two rural sites in Kilifi, Kenya, and Kiang West, the Gambia (43, 44).

We have aligned the reporting of the study methods following the standards for reporting qualitative research (SRQR) checklist (Supplementary 1) (45) and the STROBE guidelines for reporting observational studies (Supplementary 2) (46).

### 2.1 Conceptual framework

We drew on the conceptual framework of access to healthcare developed by Levesque *et al.* (47) to map the patient journey into and through the health system for people living with hypertension at six touchpoints: (1) awareness, (2) screening, (3) diagnosis, (4) treatment, (5) follow-up, and (6) blood pressure control (Figure 1) (38, 39). The framework considers demand (e.g., individual patients, their households, or communities) and supply-side (e.g., providers, institutions, or organisations) factors that shape people’s ability to access hypertension care at each touchpoint of the patient journey (8). Critical along the care pathway is an understanding of how and where people living with hypertension seek care, how they reach the providers, and how the services are obtained and used, recognising that the journey is not linear and patients can enter or exit the pathway at any stage (8).

**Figure 1.**
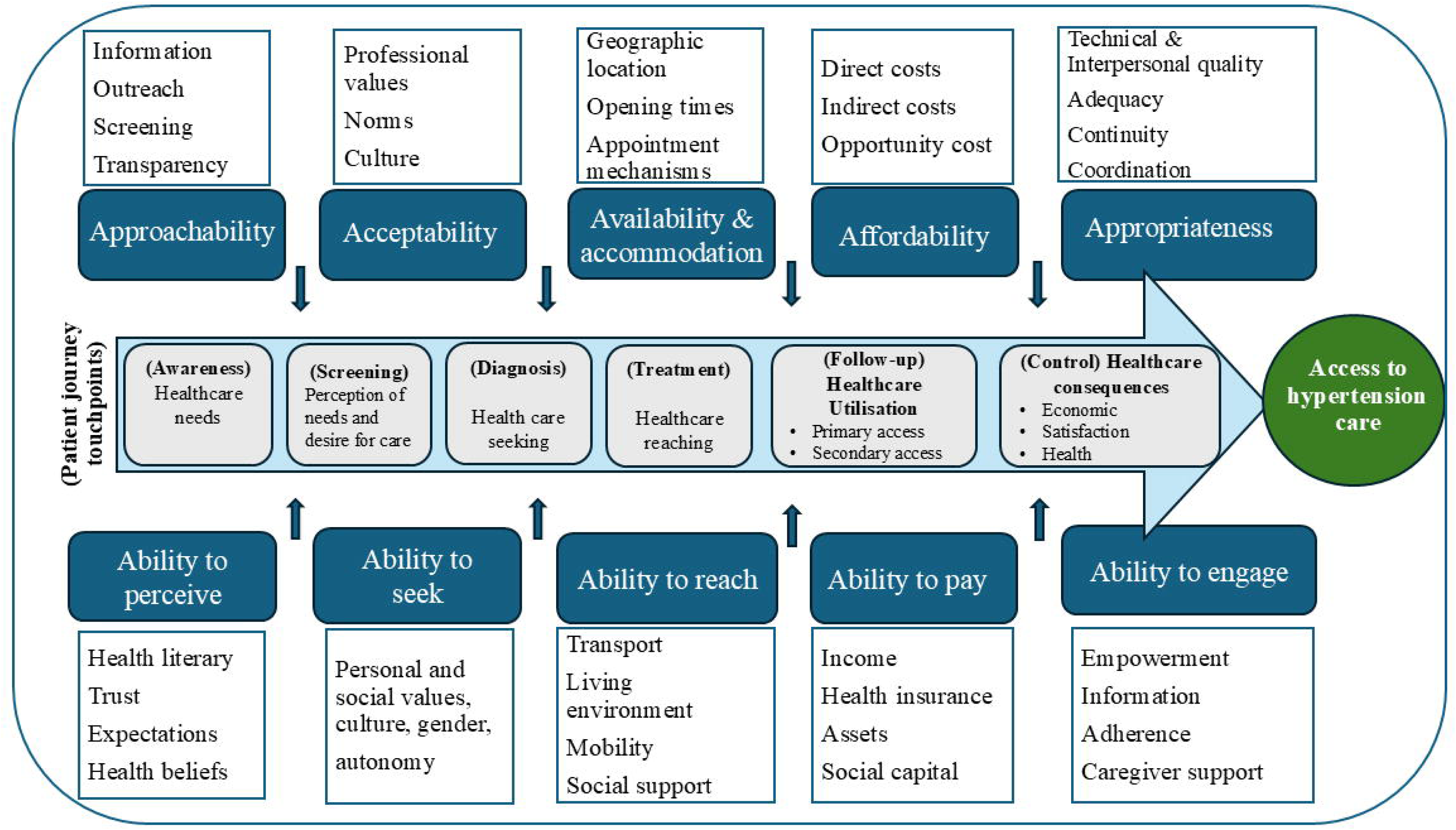
Accessibility framework along the patient journey for hypertension care (Adapted from Levesque *et al., 2013*) (47)

The six touchpoints along the patient journey are shaped by five dimensions of accessibility from a health system perspective: (1) approachability; (2) acceptability; (3) availability and accommodation; (4) affordability; and (5) appropriateness. In turn, these dimensions correspond to and interact with patients’ ability to perceive, ability to seek, ability to reach, ability to pay, and ability to engage with hypertension care services to generate access (Box 1) (47).

##### Box 1. Dimensions of accessibility

*Approachability*: the degree to which hypertension services are known to exist, information about the services is available and people needing services can reach them.

*Ability to perceive:* people at risk or those living with hypertension recognise that they need care, which is shaped by factors such as health literacy, knowledge, and beliefs.

*Acceptability:* the degree to which people living with hypertension accept healthcare services offered to them. Cultural factors and societal norms shape patients’ acceptance of healthcare services. For example, it could be that beliefs associated with systems of medicine are not acceptable to certain ethnic or religious groups.

*Ability to seek health care:* shaped by awareness and knowledge of existing health services. People living with hypertension have the power/capacity to exercise autonomy in seeking preferred care options.

*Availability and accommodation*: the extent to which hypertension services can be reached physically (e.g., physical space, availability of health workers) and in a timely manner.

*Ability to reach*: ease by which people living with hypertension can get to the services when needed.

*Affordability of care:* encompasses direct and indirect costs, including opportunity costs related to loss of income for using hypertension care services.

*Ability to pay*: patients’ capacity to mobilize resources from income, savings, sale of assets, or borrowing, to meet the costs associated with seeking hypertension care.

*Appropriateness:* extent to which the care provided meets the needs of patients with hypertension.

*Ability to engage:* level of patient involvement and participation in the care provided to them, which is shaped by, health literacy, ability to freely communicate or give feedback, and motivation to participate in care or adhere to treatment.

Source: Levesque *et al*. (47)

### 2.2 Study setting

Kenya has a devolved system of governance comprising national and 47 semiautonomous county governments (48). The national government is responsible for policy and regulatory functions as well as the management of national referral hospitals while county governments are responsible for delivering health services (49). The provision of health services across the country is shared by both public (45%) and private providers (55%), consisting of for-profit and not-for-profit providers (50). Health service delivery is organised into four tiers: community level (tier 1), primary care (tier 2), secondary referral facilities (tier 3), and tertiary referral facilities (tier 4), in order of size and complexity of services offered. Tier 1 is responsible for community-based healthcare demand creation activities as stipulated in the community health strategy; Tier 2 offers preventive and promotive services at dispensaries and health centres; Tier 3 consists of both primary (sub-county) and secondary (county) referral hospitals; and Tier 4 consists of tertiary referral hospitals under the direct management of the Ministry of Health (MoH) (51). This study was conducted in Kilifi County in the Coastal region of Kenya, a setting with a high hypertension burden. Table 1 outlines the characteristics of the study county.

**Table 1.** Key demographic and health indicators in Kilifi County.

| Indicator |  |
| --- | --- |
| Total population (2019) | 1,453,787 |
| Male | 704,089 |
| Female | 749,673 |
| Intersex | 25 |
| Share of rural population | 77% |
| Poverty rate | 46.1% |
| Health insurance coverage | 12% |
| Estimated hypertension prevalence | 26.5% |
| Health Personnel (Public sector in 2023) |  |
| Nurses per 10,000 people | 4 |
| Doctors per 10,000 people | 1 |
| Health Facilities (2022) |  |
| Dispensaries | 132 |
| Health Centres | 18 |
| Public Hospitals | 9 |
| Faith-based facilities | 25 |
| Private for-profit health facilities | 186 |

Sources: (52), (53), (54-57)

### 2.3 Study design and participant recruitment

We conducted a cross-sectional qualitative study using focus group discussions (FGDs) (n=5) and in-depth interviews (IDIs) (n=24). We used FGDs and IDIs to ensure robust and nuanced data on patient access experiences were collected from multiple perspectives thus enhancing the credibility, reliability and trustworthiness of our findings (58).

Interviews and FGDs used topic guides that were informed by the study objectives and guided by the conceptual framework. Topics sought to explore patient experiences in accessing care at each touchpoint and perceived health system-related factors that could influence access. In addition, to explore gender issues in access to hypertension care, two of the five FGDs were held separately for men and women. The interview guides were developed in English, translated into Kiswahili and Giriama, pilot-tested with five participants, reviewed, and adapted for data collection.

Interviews were conducted in person at the respective health facilities where participants were recruited. FGDs comprised between 5-8 participants and IDIs comprised 24 participants. IDIs lasted for approximately 45-60 minutes, and FGDs between 60-90 minutes. IDIs and FGDs were recorded using digital voice recorders supplemented by the interviewer’s notes. Interviews and FGDs were conducted by RO (MPH) and NK (PhD) who are both experienced qualitative and health system researchers, with the support of a trained research assistant who was fluent in the local language. Recruitment of study participants stopped after the pre-defined categories of participants had been achieved and when saturation (a point where no new substantial themes were identified with additional interviews) was attained.

### 2.4 Data collection

Data were collected between 5^th^ June 2023 and 14^th^ December 2023. Data collection was conducted in two sub-counties (Kilifi North and Kilifi South of Kilifi County), purposively selected in consultation with the county department of health officials. To achieve diverse views and experiences in accessing hypertension services, potential participants seeking care at different facility levels were approached and recruited into the study. In each sub-county, we recruited participants from one dispensary, one health centre, and the county referral hospital. Potential participants who attended scheduled clinic appointments were briefed about the study by the research team and invited to participate (with their adult caregivers where feasible and with participants’ consent) on convenient a day and time.

Participants were eligible to participate if they were 18 years or older, had a diagnosis of hypertension, and had been in care for at least six months. They were selected purposively based on their sociodemographic characteristics (e.g., age and sex), stage of the hypertension journey (newly diagnosed (i.e., ≤ 6 months), commenced treatment, with complications, those with comorbidities (i.e., hypertension and diabetes), and those who had stopped seeking care (i.e., stopped using their prescribed medication for at least three months or attending scheduled clinic appointments for at least six months). Participants who had stopped seeking care (defaulters) were recruited through household screening for hypertension as part of IHCoR’s work package two or with the help of front-line health workers (FLHWs) (43, 44). Five defaulters (two men and three women) were recruited.

### 2.5 Data analysis

IDI and FGD recordings were transcribed verbatim in Giriama and Kiswahili and then translated into English. All transcripts were reviewed for transcription accuracy by comparing them to their respective audio recordings. All transcripts were imported into the NVIvo 12 software for coding (QSR International). We used a framework approach to analyse transcripts, which involved five steps: familiarization, development of a thematic framework, coding, charting, and interpretation (59). To familiarize ourselves with the data, RO iteratively read through the interview transcripts while searching for meanings, patterns, ideas, and potential themes. NK sampled the transcripts, went through the same process independently, and consulted with RO and BT to agree on identified codes. RO then developed a coding tree, informed by the study’s conceptual framework and the ideas and themes identified in the first step. This process was reviewed and discussed with NK and BT. We then coded all transcripts based on broad themes in the patient journey touchpoints and developed subthemes identified from the data (RO, NK, BD, JB and SH). We then charted the coded data (RO), with charts checked and refined by NK and BT. In the last step (interpretation), RO examined the charted data under each thematic category to identify key concepts and the relationships between these, as well as identifying messages that are relevant to policymakers. This step was conducted in consultation with and was reviewed by all authors. Rigour and trustworthiness in the study were enhanced by a combination of 1) the use of theory, 2) methodological triangulation, 3) providing an audit trail—a detailed description of methods and analysis steps, and 4) member checking (60). Member checking was undertaken by disseminating preliminary study findings during stakeholder workshops in May and July 2024, with representatives from patient groups, community health promoters, FLHWs, and sub-national and national decision-makers. Feedback from these workshops informed us of the areas to be explored for subsequent in-depth analysis.

### 2.6 Ethical approval

Ethical approval was obtained from the KEMRI Scientific and Ethics Review Unit (Reference No. 4631), the LSHTM Ethics Committee (Reference No. 28313), and the National Committee for Science and Technology (Reference No. NACOSTI/P/23/24745). Before initiating the study procedures, we also obtained permission from the County Director of Health, sub-county health management teams, and the respective facility managers of sampled health facilities. All the participants provided written informed consent as well as their demographic data for descriptive analysis.

## 3. Results

### 3.1 General overview of participants

A total of 54 people living with hypertension and/diabetes participated in FGDs (n=30) and IDIs (n=24). Most participants were female (59%, n=32), aged between 40 and 50 years (69%, n=37), had hypertension only (63%, n=34), and were married (78%, n=42). Half (50%, n=27) of the study participants had lived with hypertension for more than five years at the time of recruitment into the study (Table 2). One potential participant who had defaulted from care declined to participate.

**Table 2.** Characteristics of FGD and IDI participants.

|  | n = 54 | % |
| --- | --- | --- |
| **Sex** |  |  |
| Male | 22 | 40.7 |
| Female | 32 | 59.3 |
| **Age (years)** |  |  |
| < 40 | 2 | 3.7 |
| 40-50 | 37 | 68.5 |
| > 50 | 15 | 27.8 |
| **NCD condition** |  |  |
| Hypertension | 34 | 63.0 |
| Hypertension + Diabetes | 20 | 37.0 |
| **Illness duration** |  |  |
| < 1 year | 5 | 9.3 |
| 1-5 years | 22 | 40.7 |
| > 5 years | 27 | 50.0 |
| **Marital status** |  |  |
| Single | 3 | 5.6 |
| Married | 42 | 77.8 |
| Widow | 7 | 12.9 |
| Divorced | 2 | 3.7 |
| **Education status** |  |  |
| No schooling | 11 | 20.4 |
| Primary | 27 | 50.0 |
| High school | 10 | 18.5 |
| Diploma/Higher | 6 | 11.1 |
| **Employment status** |  |  |
| Not employed | 29 | 53.7 |
| Informal employment | 16 | 29.6 |
| Formal employment | 9 | 16.7 |

| Family caregiver |  |  |
| --- | --- | --- |
| Yes | 11 | 20.4 |
| No | 43 | 79.6 |

We present our findings with respect to Levesque’s access framework and across the care touchpoints.

### 3.2 Approachability and ability to perceive

#### 3.2.1 Participants’ awareness of hypertension

Participants attributed hypertension to various causes such as stress because of domestic problems or financial pressures, family history, and unhealthy lifestyles. Some were aware of the risks of untreated hypertension, and they took the management of their condition seriously. This was often reported by those who had not believed their initial diagnosis or delayed the initiation of treatment for hypertension.

> *Recently, I had a complication with my kidney; I was brought here… Things were getting serious now. Then I remembered that I had been told that I have high blood pressure… I now believe that I have high blood pressure, I even adhere to medication completely* (Laughter). – Participant 2, FGD 1, Health Facility A

Lack of understanding of the causes of hypertension, and specific symptoms meant that most people were not aware of the importance of screening and seeking care. For example, there was a perception that hypertension only affected people with ‘huge’ bodies or the wealthy. Further, the lack of specific symptoms for hypertension also impaired participants’ perception on the need for screening.

> *To be sincere, when you have not had blood pressure, you cannot know whether you have it or not because you don’t know the symptoms of high blood pressure…You have to go to the hospital.* – Participant 6, FGD 1, Health Facility A

Majority of participants felt strongly that there should be better awareness education for hypertension in the community to improve hypertension knowledge.

> *People should be sensitized at the grassroots. Truly speaking, people are sick and are dying because they don’t have knowledge.* – Participant 2, FGD 2 (Female), Health Facility A

#### 3.2.2 Perceptions of approachability of screening service

Most study participants felt that hypertension screening and diagnosis services were not approachable due to lack of information on the availability of screening services. Information on hypertension was often received from health workers at diagnosis or during subsequent follow-up appointments, and sometimes from other patients.

> *I was not aware of the cause of hypertension; I did not know who to ask or who would brief me on the history of hypertension and how it starts…I did not ask anyone.* - IDI Participant 1, Health Facility B

A perceived lack of adequate information and counselling during hypertension screening and diagnosis made some participants deny their diagnosis, delay the initiation of pharmacological treatment, or discontinue their prescribed medication once they felt better. One participant commented on the lack of capacity at health facilities to screen people for hypertension and that facilities would prioritise acute care services over screening.

> *It is a challenge* [*accessing screening services*]. *For instance, there is a colleague of mine who decided to go and get screened for hypertension because he had the NHIF* [*National Health Insurance Fund*]. *When he arrived at the health facility, he was told that he was not sick as there were patients who needed urgent medical attention. However, he insisted and when he was screened, he was diagnosed with diabetes and hypertension.* – Participant 5, FGD 3 (Male), Health Facility A

### 3.3 Acceptability and ability to seek care

#### 3.3.1 Patient-provider relationship

Participants reported a diverse range of experiences that made hypertension services less acceptable to them. The failure to attain blood pressure control despite continued use of prescribed anti-hypertensives undermined acceptability of hypertension treatment as some participants abandoned their treatments. One participant related how failed blood pressure control despite medication use made formal healthcare less acceptable to her and thus, she defaulted from care. She reported:

> … *we would be given medication that would last us a month. So, I asked myself how long I would be taking the drugs, yet my blood pressure was not getting controlled. So, I decided it was better to believe in God and I continued to believe in God for five years.* – Defaulter 2 IDI Participant, Health Facility A

Similarly, other participants opted for alternative forms of treatment such as faith healing after becoming disillusioned with formal treatment.

> *Though I had started on the drugs before…I did not even tell my children when I stopped taking the drugs. Whenever they asked me, I told them that I was taking them. I was just fed up; I only took them for a week and stopped…because at church we used to be taught to have faith.* – Participant 4, FGD 1, Health Facility A

There were reports of perceived disrespectful treatment of patients by some healthcare workers which in turn reduced their willingness to seek care from their preferred health facility. One participant noted changing the health facility he attended for his routine clinic appointments to another one further away because of their experience of perceived disrespectful treatment.

> *The care at that health facility was not good…The healthcare workers used to talk rudely to patients and abuse them. If you failed to attend your scheduled appointments perhaps because of rain and went the following day, they would nearly deny you care. There was no freedom. That lady [health worker] was my daughter’s age but she behaved as though she was older than me*… – IDI Participant 2, Health Facility E

In addition, acceptability of hypertension treatment was undermined by some participants’ perceptions about the potential harm and addiction that could result from the long-term use of prescribed antihypertensive medication.

> *Some people say that once you start using antihypertensive drugs, stopping is hard. So, you will find that someone has high blood pressure but doesn’t want to use the drugs and says when he uses those drugs the disease will worsen*.…– Participant 2, FGD 2 (Female), Health Facility A

#### 3.3.2 Use of alternative forms of treatment

The use of herbal medication (commercial and traditional); either in combination with prescribed medication or singularly, was commonly reported by study participants. Overall, the use of herbal medication was occasioned by communal beliefs that one could be permanently cured of hypertension or diabetes thus enhancing the acceptability of these forms of treatment.

> *You’re told that the herbal medication is effective…so people started using herbal. The herbal people used to teach that if you take the herbal medication, hypertension would be completely healed and people like permanent healing*. – Participant 8, FGD 2 (Female), Health Facility A

However, other participants accepted hypertension treatment and were keen on ensuring they adhered to their prescribed medication. A participant shared:

> *I wasn’t aware that I had blood pressure, but when I knew, I was very focused on the drugs. Even when I came to the hospital and got only one drug and was told to buy the rest, I always bought knowing that when I leave the drugs then I will be causing a problem to myself*. – Participant 3, FGD 4, Health Facility B

Community members’ ability to seek care was undermined by fear of diagnosis of long-term conditions like hypertension, communal attitudes and perceptions towards the formal healthcare system. A participant reported:

> *People don’t want to be told that they have diabetes/hypertension…This is because when they see how you struggle with these conditions, they are very adamant about going and getting screened. When you advise people to go and get screened, they become very defensive and tell you that those are your diseases, and they don’t want to be associated with them. They deny they could have high blood pressure*. – Participant 8, FGD 2 (Female), Health Facility A

### 3.4 Availability and accommodation

#### 3.4.1 Antihypertensive medication

Persistent unavailability of antihypertensive medication across health facilities where study participants sought care was experienced as a critical barrier to accessing care by majority of respondents. A participant reported:

> *Even right now I’m worried. If I come here and I don’t get medicines, I’m worried because anything can happen when I’m at home. I wish medicines were available even for a month… Each time you come, your blood pressure reading is taken but you’re told to go and buy medicine. Now because of the constant worries and fear because of not taking medicine, your blood pressure can lead to complications*. – IDI Participant 3, Health Facility C

The unavailability of medicines eroded participants’ trust in the health system as there were perceptions of corrupt practices by FLHWs.

> *I normally wonder how government facilities lack medications, yet private chemists have them. What is that? You see. The doctor writes very fast for you to go buy medicine. God forgive me. “Go buy from so and so.” Why so and so? But as a patient, I won’t ask those questions. Where do private chemists get the anti-hypertensive medication? But we understand this is the nature of our country*…(*laughs*). – Defaulter 1 IDI Participant, Health Facility A

Participants who resided in rural areas of the county faced additional barriers in accessing medication from private pharmacies which were unavailable in rural areas. Furthermore, comorbid participants experienced challenges in accessing specific types of drug classes that were often out of stock at government facilities and not stocked at private pharmacies.

> *There is medication that is not easy to get. For example, I come from a village where there are no chemists and so, it’s* [*getting medication*] *a problem. It would be better if these medicines were available here at the hospital*. – Participant 1, FGD 3 (Male), Health Facility A

The quality of the antihypertensive medication that study participants purchased from private pharmacies was often in question as some participants reported that at times, they were given expired medicines, the wrong dosage or were given medication with a different active ingredient from the ones that they were prescribed at their clinics. A participant reported:

> *It* [*my complications*] *all started with the tablets I bought. My son took them to the hospital and showed the doctors and they told him they were not the right drugs. They had expired but I had already taken them*. – IDI Participant 4, Health Facility B

In the context of the unavailability of medications, some participants either stayed for prolonged periods without taking their medication, failed to take the required monitoring tests, or ‘shopped’ for free medication at primary healthcare facilities whereas others opted to use the relatively easily accessible alternatives like traditional herbal medication.

#### 3.4.2 Infrastructure and equipment to support access

Inadequate infrastructure and equipment to enable the provision of diagnostic and monitoring tests were reported as a barrier to access to care by study participants. For instance, due to a broken-down blood pressure monitoring machine at a primary health care facility, study participants reported that one of the health workers bought a blood pressure machine which they used to measure patients’ blood pressure at their home and charged KES 50 (USD 0.38) for the service.

In addition, study participants reported being referred to private providers due to the lack of some diagnostic and monitoring tests at public hospitals.

> *There are two tests which I was supposed to take…I came here first, then I was referred to Mombasa* [*a neighbouring city*] *because all the tests are not available here. I didn’t manage to do the tests because they required KES 45,000* [*USD 346*]. *So, we did a fundraising and went to Moshi* [*in Tanzania*]. *In Moshi, all the tests were done at a private hospital the same day and I was informed I had an enlarged heart*… – Participant 5, FGD 1, Health Facility A

Study participants who had suffered complications such as stroke experienced additional structural barriers in accessing clinics during their appointments as wheelchairs to support their movement within the health facility were either lacking or were not enough for all patients who needed such support. Such participants relied on family caregivers to coordinate and navigate the various service delivery points.

#### 3.4.3 Opening times and clinic overcrowding

Participants reported a series of linked issues that reduced the availability of services. One related to late or unpredictable clinic opening times at some facilities, which made attending scheduled clinic appointments challenging.

> *We come here in the morning and leave at around 2 or 3 pm because the healthcare worker who attends to us comes late. Sometimes she comes at 9:30 am or 10 am*…– IDI Participant 3, Health Facility C.

Some participants chose to visit other health facilities with more convenient opening times instead. A related challenge was long waiting times at health facilities along with uncertainty around the availability of needed health services. As a result, some participants discontinued their clinic attendance altogether.

Clinic overcrowding during scheduled clinic appointments was a common occurrence across all the health facilities. Study participants reported adopting various strategies to overcome this challenge. For instance, some travelled a day earlier and sought accommodation close to the hospital, others pretended to present a medical emergency to get priority medical attention while again others relied on family caregivers to come to the health facility as early as 5 am and queue for them.

Comorbid patients who were expected to attend the clinics while fasting faced additional access challenges. Whilst there were arrangements at health facilities to prioritize them during clinic visits, late/unpredictable opening hours and clinic overcrowding did not ease their access.

### 3.5 Ability to reach clinic appointments: Geographical distance and transport costs

Geographical access to health facilities was a challenge when attending routine clinic appointments, especially for participants who attended their clinics at the hospital as transport costs were a barrier to access. A participant reported:

> *You come from very far and you can’t even get money for transport. Maybe you want to come to the clinic, but you don’t have money, but you want to go…and you must buy drugs*. – Participant 7, FGD 4, Health Facility B

Whereas study participants who attended routine clinic appointments at primary health facilities had somewhat better geographical access due to the proximity of the health facilities to their homes, those who walked reported covering long distances. An elderly participant noted:

> *The challenge is that you come on foot and go back on foot. Then, there is no specific time for opening the clinic because whenever we come, we normally find the clinic not open. Therefore, you can come here in the morning and leave at around 2 or 3 pm*. – IDI Participant 3, Health Facility C

### 3.6 Affordability and ability to pay: Unaffordable hypertension services a barrier to access

Access to hypertension care was undermined by the unaffordability of services in both the public and private sectors. Of note, comorbid participants faced higher financial burdens compared to those who only had hypertension.

> *I’m taking drugs worth KES 10,000* [*USD 77*] *per month, for diabetes and hypertension. So, KES 10,000 is usually a lot. It’s more than my pension so at times I skip to save the drugs*. – Participant 8, FGD 2 (Female), Health Facility A

Uncertainty about getting needed hypertension services such as medication discouraged some study participants from attending the clinics altogether while some purchased their refill medications from private pharmacies. Similarly, due to low financial capacity, participants reported that they could not afford to adhere to the special dietary requirements to manage their condition.

> *Like the other day, I went and was prescribed drugs. When I looked at the drugs, the cost was KES 7,000* [USD 54]. *Now you see we are overwhelmed here. We are supposed to adhere to certain diets, to adhere to drugs, and with this condition, we are told not to be stressed. How will you not be stressed with everything that is before you? Now if the drugs were affordable and they are effective, things would be better*. – Participant 7, FGD 2 (Female), Health Facility A

Enrolment to the National Health Insurance Fund (NHIF) enabled access to care for participants who had the cover as they would not pay out-of-pocket for services received. However, a majority of participants did not have NHIF cover because of unaffordable monthly premiums (KES 500/ USD 4). Of note, participants who had the NHIF cover still incurred out-of-pocket payments when needed services were unavailable. In addition, there were perceptions of denial of care to NHIF-enrolled participants.

> *You see, with NHIF it depends, there are certain medicines which you are not given even if they are there*… – Participant 5, FGD 1, Health Facility A

### 3.7 Appropriateness and ability to engage: Adherence to prescribed medication and patient-provider communication

Appropriateness of care received by study participants and their ability to engage in care were undermined by interlinked provider and patient-related issues. For instance, an elderly participant reported almost dying from a medication overdose as she did not understand the instructions given to her by a FLHW. She observed:

> *I was given drugs at [name withheld] dispensary and they didn’t explain to me well that I would understand, and I took more medicine than the required dose. Ah! I almost died…they were explaining as if I knew everything, that’s why I decided to stop going there and I came here*. – IDI Participant 2, Health Facility B

Similarly, some participants failed to adhere to their medication because they did not understand the need for long-term use of drugs or perceived lack of health worker flexibility in changing their medication when they experienced adverse events. Lack of medication adherence was attributed to inadequate counselling on the long-term nature of their condition and the need for continued medication use.

> *At the private clinic they told us that once he completes his prescribed medication, he should continue doing exercise and that was all. So, we were not told that he needed the prescribed drugs for long-term management of hypertension*. – Comorbid IDI Participant 3 with Family Caregiver, Health Facility A

By contrast, other participants reported adhering to their medication without fail as well as attending routine clinic appointments as the healthcare workers had clearly communicated and counselled them well on the need for long-term medication use and clinic attendance and were thus motivated to engage in the care they received.

> *We were taught not to stop taking drugs even if our blood pressure was controlled or when someone tried to convince us otherwise, we were told not to stop taking drugs. Whoever would try to convince us otherwise would only be lying*… – IDI Participant 2, Health Facility B

Inadequate consultation time undermined some participants’ desire to engage in care. A participant noted:

> *Whom will you ask? Because while they are serving you, they are in a hurry to do other things, who will listen to you? After you have been given the drugs, they tell you to go without any explanation*. – IDI Participant 2, Health Facility B

Family caregivers were critical in enabling engagement in care for patients who had complications such as stroke, blindness or were elderly. The family caregivers coordinated how the patients got to the facility, and once there, facilitated access to care at various service delivery points. By contrast, elderly study participants who did not have family caregiver support faced additional barriers in accessing their clinics as they had challenges communicating with health workers at the health facility and did not readily comprehend their diagnosis and the medical advice given to them by health workers. A family caregiver reported:

> *When she was told that she had high blood pressure, she thought she had been diagnosed with HIV/AIDS [Giggles]…When she completed taking the two packets of her prescribed medication at that time, she believed she was healed and stopped attending clinics*. – Family Caregiver, Defaulter 5 IDI, Health Facility B

## 4. Discussion

This study aimed to examine, from the patient perspective, the demand and supply side enablers of and barriers to hypertension care at various patient journey touch points in rural Coastal Kenya. Our findings revealed several barriers at each touchpoint in access to hypertension care. One of our key findings was a lack of information on hypertension, its causes, symptoms, and the necessity for long-term treatment. This, together with participants’ underlying beliefs of hypertension not fitting with biomedical models of causes of hypertension, had a major bearing on the perceived need for screening and subsequent cascade of care for study respondents. These findings are similar to those of a recent systematic review (26) and highlight there is an urgent need for targeted awareness-raising through various mass media platforms on the value of screening for major NCDs like hypertension if the informational barriers are to be reduced or eliminated.

Previous studies have demonstrated that exposure to mass media has a significant implication on the uptake of screening services for hypertension in Kenya (17), NCDs screening uptake in Ghana (61), Indonesia (62), Ethiopia (63) and Bangladesh (64). Also, to bridge the hypertension awareness gap, measures to leverage community health systems as avenues to raise awareness through trained and well-motivated community health workers (CHWs) should be explored. There is emerging evidence from various LMIC settings that CHWs have the potential to improve knowledge, health behaviour and health outcomes related to the prevention and management of NCDs (65–68). Due to the likely concomitant increase in the demand for screening services as a result of implementing these recommendations, there is a need to strengthen the capacity of health facilities, especially at the primary healthcare level, to ensure access to hypertension screening services.

The reported perceived disrespectful treatment of patients by health workers, inadequate communication and patient counselling that undermined the appropriateness of hypertension care and participants’ ability to engage in care could have been occasioned by high patient volumes and an inadequate number of health workers that limited consultation time. Nevertheless, our finding is consistent with previous studies that have shown that gaps in provider-patient communication as well as inadequate counselling as some of the provider barriers to access to hypertension care, treatment adherence and continuity of care (8, 69–71). To overcome communication and patient counselling gaps, healthcare professionals need to be trained in providing patient-centred counselling education, especially during screening and diagnosis. This recommendation is supported by the findings of a randomized control trial that showed the effectiveness of communication skills training for physicians on health literacy skills of patients, medication adherence, and patients’ self-efficacy (i.e., patients’ confidence and ability to undertake certain tasks in managing hypertension) (72). Similarly, since provider-patient communicative interaction serves as an access and utilisation stimulus for healthcare services (73), it is also equally important to build trust with patients by ensuring open communication channels and allowing adequate consultation time during clinic visits. Strengthening supply-side factors such as ensuring an adequate number of trained health workforce, task-shifting of roles around patient education and counselling as well as strengthening oversight and supervision mechanisms of FLHWs by sub-county and county health management teams should also be strengthened. Unavailability of hypertension services was reported in this study. These were characterised by, for example, inadequate infrastructure, medication unavailability and lack of some diagnostic/monitoring tests. These findings suggest inadequate health system capacity to respond to the growing burden of NCDs and represent wider systemic issues that act as barriers to healthcare access for NCDs in LMICs (8, 26, 28, 74–77). As evidenced by our findings, participants adopted various coping strategies in the wake of these barriers. For instance, the use of alternative forms of treatment such as traditional and commercial herbal medication. Furthermore, participants lost trust in the health system due to the persistent unavailability of medication in public facilities, with some defaulting from care due to these access challenges.

Whereas there are no straightforward solutions to these health system-wide access challenges, various short and long-term strategies could be implemented by FLHWs, county governments and the MoH to enhance access to care. In the short term, FLHWs should be trained to implement culturally sensitive patient empowerment interventions on the effectiveness of prescribed medication in ensuring blood pressure control compared with herbal medication, thus building trust and improving patient engagement with mainstream services (78). Further, as shown in our results and elsewhere (26, 28, 74, 77), measures to involve family caregivers in the delivery of hypertension care would be critical in enabling access to care, especially for elderly patients or those with complications. In the long term, county governments should increasingly prioritise investments in the health sector, particularly increasing budgetary allocations to NCDs to ensure consistent availability of medicine, equipment and infrastructure for hypertension care. Also, the MoH should implement new and strengthen existing regulatory frameworks for private pharmacies, including the licensing of commercial herbal medicine practitioners through government agencies such as the Pharmacy and Poisons Board to ensure the sale of quality and efficacious antihypertensive medicines.

Unaffordability of care and patients’ limited capacity to pay was an access barrier to hypertension treatment and attendance of routine clinic appointments. Nevertheless, having NHIF cover was an access enabler for participants who had the cover. Previous studies conducted in Kenya have shown that costs for NCD care are mainly driven by medication (18, 79) and transport costs (18, 79), with screening, diagnosis and treatment costs being significantly higher in the private relative to the public sector (80). Unaffordability of hypertension care services is occasioned by inadequate prepayment mechanisms in the Kenyan health system. For example, recent studies that evaluated the responsiveness of the NHIF to NCD patient needs as well as its effectiveness in cushioning households from incurring catastrophic health expenditures (CHE) found that whereas NHIF expanded access to NCD care, a narrow range of services were covered and households that were enrolled in NHIF were not better protected from incurring CHE compared with households without NHIF, with a high attrition rate (76.7%) after one-year follow-up, suggesting unaffordability of the monthly NHIF premiums (36, 37). The ongoing health financing reforms in Kenya provide a window of opportunity to include this study’s findings into account while designing the benefit package of the Emergency, Accidents and Chronic Illness Fund. Also, as has been demonstrated by studies from Ethiopia (81), South Africa (82), and the Philippines (83), there is a multi-pronged benefit of increasing excise taxes on potentially harmful health products such as tobacco, alcohol and sugar-sweetened beverages that are modifiable risk factors for NCDs. Implementing excise tax increments on such products has been shown to generate additional revenues. These revenues could increase the resource envelope for the health sector in tackling NCDs. Importantly, such excise tax hikes have also averted CHE and improved health outcomes, with the poor benefitting most (84, 85). Future studies should explore the potential distributional impact of instituting such excise tax increments in Kenya.

### 4.1 Study strengths and reflections on Levesque’s framework of access

To our knowledge, this is the first qualitative study to explore, from the patient perspective, demand and supply enablers of and barriers to access to hypertension care in a rural setting in Kenya. A key strength of this study is the use of Levesque’s conceptualization of access which allowed for an in-depth analysis of a complex and dynamic process of access at the population and health system interfaces. This conceptualisation of access helps move from critiquing the supply side to recognizing interactions between characteristics related to individuals and health systems, therefore enabling the formulation of recommendations to address both (not just health systems).

A scoping review that aimed to identify and analyse empirical studies that have applied Levesque’s conceptual framework of access and document the successes and challenges in its use recommended the inclusion of time-related elements of access (e.g., travel time) (86). In our study, however, we were able to analyse time-related elements of access under the availability and accommodation dimension of access and the corresponding ability to reach. We therefore feel that this concern may not be relevant to our study.

Nevertheless, we concur that some dimensions of access and corresponding abilities in the Levesque framework could have more than one application (86). For instance, health facilities that are located far from people’s homes would be interpreted under geographical location if one looks at it from the availability dimension. By contrast, if the cost of transportation is considered, direct costs under the affordability dimension would be considered instead (86). Unlike previous studies that have only used some dimensions and abilities of access, our study applied Levesque’s framework in its entirety thus minimizing this challenge. The second strength of our study is the exploration of access enablers and barriers using more than one method of data collection: FGDs and IDIs. This allowed for methodological rigour and triangulation of findings from multiple perspectives, thus ensuring the credibility and reliability of our findings. Further, the trustworthiness of our findings was substantiated by providing a research audit trail and member checking during stakeholder workshops where preliminary findings were disseminated. Third, we explored access to hypertension care along various patient journey touchpoints and thus presented a comprehensive assessment of access to hypertension care.

### 4.2 Study limitations

The findings of this study should be interpreted in light of the following limitations. First, whereas attempts were made to recruit a broad range of participants, including those who had defaulted from care, only five participants (two men and three women) were recruited. As a result, our findings under-represent access experiences from patients who have disengaged from the formal health sector. Second, gender has been documented as a determinant of access to healthcare in the literature (26, 77). Whereas we did not set out to explicitly explore gender and access to hypertension care, we held two separate FGDs with men and women in a bid to assess gender barriers/enablers to access along the patient journey. We did not find gendered issues around access to hypertension care. Given these limitations, we recommend that future studies should 1) conduct a comprehensive assessment of experiences from individuals who have disengaged from formal care, and 2) explore gender issues in access to NCD care in Kenya.

### 4.3 Conclusions

The findings from our study present a critical mix of barriers to and enablers of access to hypertension care at the individual, community and health system level in a rural context in Kenya’s health system. We also offer potential policy recommendations to reduce or eliminate the barriers. Comprehensive patient counselling at screening and diagnosis, targeted awareness raising on hypertension through CHWs and various mass media platforms, mechanisms to monitor and oversight FLHWs as well as private pharmacies, including herbal medicine practitioners and prioritisation of NCDs through increased budgetary allocations to ensure availability and affordability of hypertension service should be prioritised.

## Supporting information

SRQR and STROBE checklists

## Data Availability

The data presented in this study are available upon reasonable request from the corresponding author through the. Public deposition of the transcripts would breach compliance with the approved protocol. During ethical approval, the transcripts were stated would be available publicly upon reasonable request through KEMRI-Wellcome Trust’s data governance committee through the. This was because at the time of data collection, it was deemed a sensitive topic, it was paramount not to compromise patient privacy, and allow free participation among the respondents. We are therefore kindly requesting for this exemption.

## Author Contributions

Benjamin Tsofa, Ellen Nolte, Edwine Barasa, Samson Muchina Kinyanjui, Anthony Etyang, Pablo Perel: Conceptualization, Funding Acquisition, Investigation, Methodology, Validation, Supervision, Review and Editing.

Robinson Oyando: Conceptualization, Project Administration, Investigation, Methodology, Data Collection, Data Curation, Formal Analysis, Original Draft Preparation. Nancy Kagwanja: Project Administration, Investigation, Methodology, Data Collection, Data Curation, Validation, Review and Editing.

Brahima Diallo, Syreen Hassan, Jainaba Badjie, Ruth Lucinde, Noni Mumba: Investigation, Data Curation, Validation, Resources, Review and Editing.

IHCoR-Africa Collaborators: Conceptualization, Investigation, Validation.

## Acknowledgements

We acknowledge the support of John Koi during data collection and thank study participants for sharing their experiences with the research team. We thank Ruth Willis (LSHTM) and Rose Muthuri (KWTRP) for their insightful comments on an earlier draft of the paper.

## References

1. World Health Organization. Global report on hypertension: the race against a silent killer: World Health Organization; 2023.

2. Vos T, Lim SS, Abbafati C, Abbas KM, Abbasi M, Abbasifard M, et al. Global burden of 369 diseases and injuries in 204 countries and territories, 1990–2019: a systematic analysis for the Global Burden of Disease Study 2019. The lancet. 2020;396(10258):1204–22.

3. IHME. High Systolic Blood Pressure-Level 2 Risk. IHME, University of Washington Seattle, WA, USA; 2019.

4. Murray CJ, Aravkin AY, Zheng P, Abbafati C, Abbas KM, Abbasi-Kangevari M, et al. Global burden of 87 risk factors in 204 countries and territories, 1990–2019: a systematic analysis for the Global Burden of Disease Study 2019. The lancet. 2020;396(10258):1223–49.

5. Zhou B, Carrillo-Larco RM, Danaei G, Riley LM, Paciorek CJ, Stevens GA, et al. Worldwide trends in hypertension prevalence and progress in treatment and control from 1990 to 2019: a pooled analysis of 1201 population-representative studies with 104 million participants. The Lancet. 2021;398(10304):957–80.

6. Schutte AE, Srinivasapura Venkateshmurthy N, Mohan S, Prabhakaran D. Hypertension in low-and middle-income countries. Circulation research. 2021;128(7):808–26.

7. Stein DT, Reitsma MB, Geldsetzer P, Agoudavi K, Aryal KK, Bahendeka S, et al. Hypertension care cascades and reducing inequities in cardiovascular disease in low-and middle-income countries. Nature Medicine. 2024;30(2):414–23.

8. Legido-Quigley H, Naheed A, De Silva HA, Jehan I, Haldane V, Cobb B, et al. Patients’ experiences on accessing health care services for management of hypertension in rural Bangladesh, Pakistan and Sri Lanka: a qualitative study. PloS one. 2019;14(1):e0211100.

9. Geldsetzer P, Manne-Goehler J, Marcus M-E, Ebert C, Zhumadilov Z, Wesseh CS, et al. The state of hypertension care in 44 low-income and middle-income countries: a cross-sectional study of nationally representative individual-level data from 1· 1 million adults. The Lancet. 2019;394(10199):652–62.

10. Kayima J, Wanyenze RK, Katamba A, Leontsini E, Nuwaha F. Hypertension awareness, treatment and control in Africa: a systematic review. BMC cardiovascular disorders. 2013;13:1–11.

11. World Bank. Rural population (% of total population) - Sub-Saharan Africa. Available at: https://data.worldbank.org/indicator/SP.RUR.TOTL.ZS?locations=ZG (Accessed on 01 August 2024) 2023 [

12. Karinja M, Pillai G, Schlienger R, Tanner M, Ogutu B. Care-seeking dynamics among patients with diabetes mellitus and hypertension in selected rural settings in Kenya. International journal of environmental research and public health. 2019;16(11):2016.

13. Zhou B, Bentham J, Di Cesare M, Bixby H, Danaei G, Cowan MJ, et al. Worldwide trends in blood pressure from 1975 to 2015: a pooled analysis of 1479 population-based measurement studies with 19· 1 million participants. The Lancet. 2017;389(10064):37–55.

14. World Health Organization. Hypertension Kenya 2023 country profile. Available at: https://www.who.int/publications/m/item/hypertension-ken-2023-country-profile (Accessed on 23/1/2025) 2023 [

15. Mohamed SF, Mutua MK, Wamai R, Wekesah F, Haregu T, Juma P, et al. Prevalence, awareness, treatment and control of hypertension and their determinants: results from a national survey in Kenya. BMC public health. 2018;18:1–10.

16. Gnugesser E, Chwila C, Brenner S, Deckert A, Dambach P, Steinert J, et al. The economic burden of treating uncomplicated hypertension in Sub-Saharan Africa: a systematic literature review. BMC Public Health. 2022;22(1):1507.

17. Oyando R, Barasa E, Ataguba JE. Socioeconomic inequity in the screening and treatment of hypertension in Kenya: evidence from a National Survey. Frontiers in Health Services. 2022;2:786098.

18. Oyando R, Njoroge M, Nguhiu P, Kirui F, Mbui J, Sigilai A, et al. Patient costs of hypertension care in public health care facilities in Kenya. The International journal of health planning and management. 2019;34(2):e1166–e78.

19. Palafox B, McKee M, Balabanova D, AlHabib KF, Avezum AJ, Bahonar A, et al. Wealth and cardiovascular health: a cross-sectional study of wealth-related inequalities in the awareness, treatment and control of hypertension in high-, middle-and low-income countries. International journal for equity in health. 2016;15:1–14.

20. Brundisini F, Giacomini M, DeJean D, Vanstone M, Winsor S, Smith A. Chronic disease patients’ experiences with accessing health care in rural and remote areas: a systematic review and qualitative meta-synthesis. Ontario health technology assessment series. 2013;13(15):1.

21. Mogaka JN, Sharma M, Temu T, Masyuko S, Kinuthia J, Osoti A, et al. Prevalence and factors associated with hypertension among adults with and without HIV in Western Kenya. Plos one. 2022;17(1):e0262400.

22. Rockers PC, Laing RO, Wirtz VJ. Equity in access to non-communicable disease medicines: a cross-sectional study in Kenya. BMJ Global Health. 2018;3(3):e000828.

23. Herbst AG, Olds P, Nuwagaba G, Okello S, Haberer J. Patient experiences and perspectives on hypertension at a major referral hospital in rural southwestern Uganda: a qualitative analysis. BMJ open. 2021;11(1):e040650.

24. Lynch HM, Green AS, Clarke Nanyonga R, Gadikota-Klumpers DD, Squires A, Schwartz JI, et al. Exploring patient experiences with and attitudes towards hypertension at a private hospital in Uganda: a qualitative study. International journal for equity in health. 2019;18:1–7.

25. Naanyu V, Vedanthan R, Kamano JH, Rotich JK, Lagat KK, Kiptoo P, et al. Barriers influencing linkage to hypertension care in Kenya: qualitative analysis from the LARK hypertension study. Journal of general internal medicine. 2016;31:304–14.

26. Brathwaite R, Hutchinson E, McKee M, Palafox B, Balabanova D. The long and winding road: a systematic literature review conceptualising pathways for hypertension care and control in low-and middle-income countries. Int J Health Policy Manag. 2022;11(3):257.

27. Sorato MM, Davari M, Kebriaeezadeh A, Sarrafzadegan N, Shibru T, Fatemi B. Reasons for poor blood pressure control in eastern sub-Saharan Africa: looking into 4P’s (primary care, professional, patient, and public health policy) for improving blood pressure control: a scoping review. BMC cardiovascular disorders. 2021;21:1–15.

28. Amarteyfio KNAA, Bondzie EPK, Reichenberger V, Afun NEE, Cofie AKM, Agyekum MP, et al. Factors influencing access, quality and utilisation of primary healthcare for patients living with hypertension in West Africa: a scoping review. BMJ Open. 2024;14(12):e088718.

29. Gafane-Matemane LF, Craig A, Kruger R, Alaofin OS, Ware LJ, Jones ES, et al. Hypertension in sub-Saharan Africa: the current profile, recent advances, gaps, and priorities. Journal of Human Hypertension. 2024:1–16.

30. Kabir A, Karim MN, Islam RM, Romero L, Billah B. Health system readiness for non-communicable diseases at the primary care level: a systematic review. BMJ open. 2022;12(2):e060387.

31. Onyango MA, Vian T, Hirsch I, Salvi DD, Laing R, Rockers PC, et al. Perceptions of Kenyan adults on access to medicines for non-communicable diseases: A qualitative study. PLoS One. 2018;13(8):e0201917.

32. Cárdenas MK, Pérez-León S, Singh SB, Madede T, Munguambe S, Govo V, et al. Forty years after Alma-Ata: primary health-care preparedness for chronic diseases in Mozambique, Nepal and Peru. Global health action. 2021;14(1):1975920.

33. Williams KN, Tenorio-Mucha J, Campos-Blanco K, Underhill LJ, Valdés-Velásquez A, Herbozo AF, et al. Health system barriers to hypertension care in Peru: Rapid assessment to inform organizational-level change. PLOS Global Public Health. 2024;4(8):e0002404.

34. Heller O, Somerville C, Suggs LS, Lachat S, Piper J, Aya Pastrana N, et al. The process of prioritization of non-communicable diseases in the global health policy arena. Health policy and planning. 2019;34(5):370–83.

35. Luna F, Luyckx VA. Why have non-communicable diseases been left behind? Asian bioethics review. 2020;12(1):5–25.

36. Oyando R, Were V, Koros H, Mugo R, Kamano J, Etyang A, et al. Evaluating the effectiveness of the National Health Insurance Fund in providing financial protection to households with hypertension and diabetes patients in Kenya. International journal for equity in health. 2023;22(1):107.

37. Oyando R, Were V, Willis R, Koros H, Kamano JH, Naanyu V, et al. Examining the responsiveness of the National Health Insurance Fund to people living with hypertension and diabetes in Kenya: a qualitative study. BMJ open. 2023;13(7):e069330.

38. Devi R, Kanitkar K, Narendhar R, Sehmi K, Subramaniam K. A narrative review of the patient journey through the lens of non-communicable diseases in low-and middle-income countries. Advances in Therapy. 2020;37(12):4808–30.

39. Bharatan T, Devi R, Huang P-H, Javed A, Jeffers B, Lansberg P, et al. A methodology for mapping the patient journey for noncommunicable diseases in low-and middle-income countries. Journal of healthcare leadership. 2021:35–46.

40. Naanyu V, Willis R, Kamano J, Koros H, Murphy A, Perel P, et al. Managing diabetes and hypertension in western Kenya: A qualitative study of experiences of patients supported by the primary health integrated care for chronic conditions (PIC4C) model of care. PLOS Global Public Health. 2024;4(8):e0003245.

41. Ng G, Raskin E, Wirtz VJ, Banks KP, Laing RO, Kiragu ZW, et al. Coping with access barriers to non-communicable disease medicines: qualitative patient interviews in eight counties in Kenya. BMC Health Services Research. 2021;21:1–10.

42. Golembiewski EH, Gravholt DL, Roldan VDT, Naranjo EPL, Vallejo S, Bautista AG, et al. Rural patient experiences of accessing care for chronic conditions: a systematic review and thematic synthesis of qualitative studies. The Annals of Family Medicine. 2022;20(3):266–72.

43. Perkins AD, Awori JO, Jobe M, Lucinde RK, Siemonsma M, Oyando R, et al. Determining the optimal diagnostic and risk stratification approaches for people with hypertension in two rural populations in Kenya and The Gambia: a study protocol for IHCoR-Africa Work Package 2. NIHR Open Research. 2023;3.

44. Diallo BA, Hassan S, Kagwanja N, Oyando R, Badjie J, Mumba N, et al. Managing hypertension in rural Gambia and Kenya: Protocol for a qualitative study exploring the experiences of patients, health care workers, and decision-makers. NIHR Open Research. 2024;4:5.

45. O’Brien BC, Harris IB, Beckman TJ, Reed DA, Cook DA. Standards for reporting qualitative research: a synthesis of recommendations. Academic medicine. 2014;89(9):1245–51.

46. Von Elm E, Altman DG, Egger M, Pocock SJ, Gøtzsche PC, Vandenbroucke JP. The Strengthening the Reporting of Observational Studies in Epidemiology (STROBE) statement: guidelines for reporting observational studies. The lancet. 2007;370(9596):1453-7.

47. Levesque J-F, Harris MF, Russell G. Patient-centred access to health care: conceptualising access at the interface of health systems and populations. International journal for equity in health. 2013;12:1–9.

48. Government of Kenya. The Constitution of Kenya. Nairobi; 2010.

49. Minsitry of Health. Kenya Health Policy 2014-2030: Towards attaining the highest standard of health. Nairobi; 2014.

50. Moturi AK, Suiyanka L, Mumo E, Snow RW, Okiro EA, Macharia PM. Geographic accessibility to public and private health facilities in Kenya in 2021: an updated geocoded inventory and spatial analysis. Frontiers in Public Health. 2022;10:1002975.

51. Ministry of Health. Kenya health policy 2014-2030: Ministry of Health; 2014.

52. KNBS. Kenya population and housing census. Nairobi: Kenya National Bureau of Statistics. 2019.

53. County Government of Kilifi. County Integrated Development Plan 2023-2027: Accelerating Socioeconomic Transfromation for Inclusive Growth. 2023.

54. Tsofa B OL, Ngala T, et al.,. Kilifi county health sector review. In: Government KC,. Kilifi: Kilifi County Government; 2023.

55. Ministry of Health. Kenya Health Information System: https://hiskenya.org/dhis-web-commons/security/login.action#/data-set-report (Accessed on 21 August 2024) 2024 [

56. Etyang AO, Warne B, Kapesa S, Munge K, Bauni E, Cruickshank JK, et al. Clinical and epidemiological implications of 24-hour ambulatory blood pressure monitoring for the diagnosis of hypertension in Kenyan adults: a Population-Based Study. Journal of the American Heart Association. 2016;5(12):e004797.

57. Kenya National Bureau of Statistics (KNBS) and ICF. Kenya Demographic and Health Survey 2022 Nairobi, Kenya, and Rockville, Maryland, USA: KNBS and ICF; 2023.

58. Lambert SD, Loiselle CG. Combining individual interviews and focus groups to enhance data richness. Journal of advanced nursing. 2008;62(2):228–37.

59. Ritchie J, Spencer L. Qualitative data analysis for applied policy research. Analyzing qualitative data: Routledge; 2002. p. 173–94.

60. Gilson L, Organization WH. Health policy and system research: a methodology reader: the abridged version: World Health Organization; 2013.

61. Konkor I, Bisung E, Soliku O, Ayanore M, Kuuire V. Exposure to mass media chronic health campaign messages and the uptake of non-communicable disease screening in Ghana. Plos one. 2024;19(5):e0302942.

62. Firdaus R, Dewi YLR, Murti B. Factors Affecting the Uptake of Non Communicable Disease Screening Service: A Multiple Logistic Regression in Kapuas Hulu, West Kalimantan. Journal of Epidemiology and Public Health. 2019;4(4):373–85.

63. Debebe H, Ketema B, Rossner SS, Negash S, Addissie A, Kaba M, et al. The Promotion of Non-Communicable Disease Screening in Gurage Zone, Ethiopia: A Mixed-Method Study. Diseases. 2024;12(11):294.

64. Nessa A, Hussain MA, Ur Rashid MH, Akhter N, Roy JS, Afroz R. Role of print and audiovisual media in cervical cancer prevention in Bangladesh. Asian Pacific Journal of Cancer Prevention. 2013;14(5):3131–7.

65. Alaofè H, Asaolu I, Ehiri J, Moretz H, Asuzu C, Balogun M, et al. Community health workers in diabetes prevention and management in developing countries. Annals of global health. 2017;83(3-4):661–75.

66. Jeet G, Thakur J, Prinja S, Singh M. Community health workers for non-communicable diseases prevention and control in developing countries: evidence and implications. PloS one. 2017;12(7):e0180640.

67. Mbuthia GW, Mwangi J, Magutah K, Oguta JO, Ngure K, McGarvey ST. Preliminary efficacy of a community health worker homebased intervention for the control and management of hypertension in Kiambu County, Kenya-a randomized control trial. Plos one. 2024;19(8):e0293791.

68. Doresha L-M, Mash R. The role of community health workers in non-communicable diseases in Cape Town, South Africa: descriptive exploratory qualitative study. BMC Primary Care. 2024;25(1):176.

69. Shrestha S, Shrestha A, Koju RP, LoGerfo JP, Karmacharya BM, Sotoodehnia N, et al. Barriers and facilitators to treatment among patients with newly diagnosed hypertension in Nepal. Heart Asia. 2018;10(2).

70. Khatib R, Schwalm J-D, Yusuf S, Haynes RB, McKee M, Khan M, et al. Patient and healthcare provider barriers to hypertension awareness, treatment and follow up: a systematic review and meta-analysis of qualitative and quantitative studies. PloS one. 2014;9(1):e84238.

71. Gabert R, Ng M, Sogarwal R, Bryant M, Deepu R, McNellan CR, et al. Identifying gaps in the continuum of care for hypertension and diabetes in two Indian communities. BMC health services research. 2017;17:1–11.

72. Tavakoly Sany SB, Behzhad F, Ferns G, Peyman N. Communication skills training for physicians improves health literacy and medical outcomes among patients with hypertension: a randomized controlled trial. BMC health services research. 2020;20:1–10.

73. Thiede M. Information and access to health care: is there a role for trust? Social science & medicine. 2005;61(7):1452–62.

74. Dhungana RR, Pedisic Z, Pandey AR, Shrestha N, de Courten M. Barriers, enablers and strategies for the treatment and control of hypertension in Nepal: a systematic review. Frontiers in Cardiovascular Medicine. 2021;8:716080.

75. Albelbeisi AH, Albelbeisi A, El Bilbeisi AH, Taleb M, Takian A, Akbari-Sari A. Public sector capacity to prevent and control of noncommunicable diseases in twelve low-and middle-income countries based on WHO-PEN standards: a systematic review. Health Services Insights. 2021;14:1178632920986233.

76. Ammoun R, Wami WM, Otieno P, Schultsz C, Kyobutungi C, Asiki G. Readiness of health facilities to deliver non-communicable diseases services in Kenya: a national cross-sectional survey. BMC health services research. 2022;22(1):1–11.

77. Dawkins B, Renwick C, Ensor T, Shinkins B, Jayne D, Meads D. What factors affect patients’ ability to access healthcare? An overview of systematic reviews. Tropical Medicine & International Health. 2021;26(10):1177–88.

78. Palafox B, Balabanova D, Loreche AM, Mat-Nasir N, Ariffin F, Md-Yasin M, et al. Pathways to Hypertension Control: Unfinished Journeys of Low-Income Individuals in Malaysia and the Philippines. The International Journal of Health Planning and Management. 2024.

79. Oyando R, Njoroge M, Nguhiu P, Sigilai A, Kirui F, Mbui J, et al. Patient costs of diabetes mellitus care in public health care facilities in Kenya. The International journal of health planning and management. 2020;35(1):290–308.

80. Subramanian S, Gakunga R, Kibachio J, Gathecha G, Edwards P, Ogola E, et al. Cost and affordability of non-communicable disease screening, diagnosis and treatment in Kenya: Patient payments in the private and public sectors. PloS one. 2018;13(1):e0190113.

81. Chakrabarti A, Memirie ST, Yigletu S, Mirutse MK, Verguet S. The potential distributional health and financial benefits of increased tobacco taxes in Ethiopia: Findings from a modeling study. SSM-Population Health. 2022;18:101097.

82. Boachie MK, Hofman K, Goldstein S, Thsehla E. Modelling the potential impact of a tax on fruit juice in South Africa: implications for the primary prevention of type 2 diabetes and health financing. BMC nutrition. 2024;10(1):145.

83. Saxena A, Koon AD, Lagrada-Rombaua L, Angeles-Agdeppa I, Johns B, Capanzana M. Modelling the impact of a tax on sweetened beverages in the Philippines: an extended cost–effectiveness analysis. Bulletin of the World Health Organization. 2019;97(2):97.

84. Chaloupka FJ, Powell LM, Warner KE. The use of excise taxes to reduce tobacco, alcohol, and sugary beverage consumption. Annual review of public health. 2019;40(1):187–201.

85. Miracolo A, Sophiea M, Mills M, Kanavos P. Sin taxes and their effect on consumption, revenue generation and health improvement: a systematic literature review in Latin America. Health Policy and Planning. 2021;36(5):790–810.

86. Cu A, Meister S, Lefebvre B, Ridde V. Assessing healthcare access using the Levesque’s conceptual framework–a scoping review. International journal for equity in health. 2021;20(1):116.

