## Supplementary material for "Access to hypertension services and health-seeking experiences in rural Coastal Kenya: A qualitative study": SRQR and STROBE checklists: S1_SRQR_IHCoR.pdf

| Topic | No. | Item | Page No |
|---|---|---|---|
| **Title** | S1 | Concise description of the nature and topic of the study Identifying the study as qualitative or indicating the approach (e.g., ethnography, grounded theory) or data collection methods (e.g., interview, focus group) is recommended | Pg. 1, Line 1-2 |
| **Abstract** | S2 | Summary of key elements of the study using the abstract format of the intended publication; typically includes background, purpose, methods, results, and conclusions | Pg. 2, Line 31-64 |
| **Introduction** | | | |
| Problem formulation | S3 | Description and significance of the problem/phenomenon studied; review of relevant theory and empirical work; problem statement | Pg.4-6, Line 71-136 |
| Purpose or research question | S4 | Purpose of the study and specific objectives or questions | Pg. 6, Line 119-136 |
| **Methods** | | | |
| Qualitative approach and research paradigm | S5 | Qualitative approach (e.g., ethnography, grounded theory, case study, phenomenology, narrative research) and guiding theory if appropriate; identifying the research paradigm (e.g., postpositivist, constructivist/ interpretivist) is also recommended; rationale | Pg.7-9, Line 152-202 |
| Researcher characteristics and reflexivity | S6 | Researchers' characteristics that may influence the research, including personal attributes, qualifications/experience, relationship with participants, assumptions, and/or presuppositions; potential or actual interaction between researchers' characteristics and the research questions, approach, methods, results, and/or transferability | Pg. 11, Line 239-242 |
| Context | S7 | Setting/site and salient contextual factors; rationale | Pg.9, Line 206-222 |
| Sampling strategy | S8 | How and why research participants, documents, or events were selected; criteria for deciding when no further sampling was necessary (e.g., sampling saturation); rationale | Page 11; Line 230-245 |
| Ethical issues pertaining to human subjects | S9 | Documentation of approval by an appropriate ethics review board and participant consent, or explanation for lack thereof; other confidentiality and data security issues | Pg. 13, Line 296-302 |

| | | | |
|---|---|---|---|
| Data collection methods | S10 | Types of data collected; details of data collection procedures including (as appropriate) start and stop dates of data collection and analysis, iterative process, triangulation of sources/methods, and modification of procedures in response to evolving study findings; rationale | Pg. 11-12; Line 240-258 |
| Data collection instruments and technologies | S11 | Description of instruments (e.g., interview guides, questionnaires) and devices (e.g., audio recorders) used for data collection; if/how the instrument(s) changed over the course of the study | Pg.11-12, Line 248-266 |
| Units of study | S12 | Number and relevant characteristics of participants, documents, or events included in the study; level of participation (could be reported in results) | Pg.15-16, Line 305-312 |
| Data processing | S13 | Methods for processing data prior to and during analysis, including transcription, data entry, data management and security, verification of data integrity, data coding, and anonymization/deidentification of excerpts | Pg. 12, Line 261-264 |
| Data analysis | S14 | Process by which inferences, themes, etc., were identified and developed, including the researchers involved in data analysis; usually references a specific paradigm or approach; rationale | Pg. 12-13, Line 269-293 |
| Techniques to enhance trustworthiness | S15 | Techniques to enhance trustworthiness and credibility of data analysis (e.g., member checking, audit trail, triangulation); rationale | Pg. 13, Line 286-293 |
| **Results** | | | |
| Synthesis and interpretation | S16 | Main findings (e.g., interpretations, inferences, and themes); might include development of a theory or model, or integration with prior research or theory | Pg. 16-28, Line 316-637 |
| Links to empirical data | S17 | Evidence (e.g., quotes, field notes, text excerpts, photographs) to substantiate analytic findings | Pg. 16-29, Line 316-637 |
| **Discussion** | | | |
| Integration with prior work, implications, transferability, and contribution(s) to the field | S18 | Short summary of main findings; explanation of how findings and conclusions connect to, support, elaborate on, or challenge conclusions of earlier scholarship; discussion of scope of application/ generalizability; identification of unique contribution(s) to scholarship in a discipline or field | Pg. 30-35, Line 641-771; |

| | | | |
|---|---|---|---|
| Limitations | S19 | Trustworthiness and limitations of findings | Pg. 35, Line 773-784 |
| **Other** | | | |
| Conflicts of interest | S20 | Potential sources of influence or perceived influence on study conduct and conclusions; how these were managed | Pg. 37, Line 823 |
| Funding | S21 | Sources of funding and other support; role of funders in data collection, interpretation, and reporting | Pg. 36, Line 810-815 |

**Reference**

[1] O'Brien BC, Harris IB, Beckman TJ, Reed DA, Cook DA. Standards for reporting qualitative research: a synthesis of recommendations. Acad Med. 2014;89(9):1245-1251.
