## Supplementary material for "Access to hypertension services and health-seeking experiences in rural Coastal Kenya: A qualitative study": SRQR and STROBE checklists: S2_STROBE_IHCoR.pdf

STROBE Statement—checklist of items that should be included in reports of observational studies

| | Item No. | Recommendation | Page No. | Relevant text from manuscript |
|---|---|---|---|---|
| **Title and abstract** | 1 | (*a*) Indicate the study's design with a commonly used term in the title or the abstract | 1,2 | '*a qualitative study'* (full title line 2, abstract line 36-37) |
| | | (*b*) Provide in the abstract an informative and balanced summary of what was done and what was found | 2,3 | lines 36-58 |
| **Introduction** | | | | |
| Background/rationale | 2 | Explain the scientific background and rationale for the investigation being reported | 4-6 | lines 71-132 |
| Objectives | 3 | State specific objectives, including any prespecified hypotheses | 6 | lines 132-136 |
| **Methods** | | | | |
| Study design | 4 | Present key elements of study design early in the paper | 10 | lines 225 |
| Setting | 5 | Describe the setting, locations, and relevant dates, including periods of recruitment, exposure, follow-up, and data collection | 9-11 | Setting and location: lines 206-222<br>Date reporting:<br>- Recruitment line 236-245,<br>- Data collection period lines 248, as appropriate for qualitative study reporting (SRQR Checklist item 10)<br>- The concept of 'exposure' is not appropriate for this study design |
| Participants | 6 | (*a*) *Cohort study*—Give the eligibility criteria, and the sources and methods of selection of participants. Describe methods of follow-up<br>*Case-control study*—Give the eligibility criteria, and the sources and methods of case ascertainment and control selection. Give the rationale for the choice of cases and controls<br>*Cross-sectional study*—Give the eligibility criteria, and the sources and methods of selection of participants | 10 | Lines 257-263 explain eligibility criteria '*Participants were eligible to participate if they were 18 years or older, had a diagnosis of hypertension, and had been in care for at least six months'*. Lines 250-258 describe the purposive sampling strategy (SRQR Checklist item 8), as appropriate for the qualitative study design. |
| | | (*b*) *Cohort study*—For matched studies, give matching criteria and number of exposed and unexposed | | Not applicable to this study design |

| | | | | |
|---|---|---|---|---|
| | | *Case-control study*—For matched studies, give matching criteria and the number of controls per case | | |
| Variables | 7 | Clearly define all outcomes, exposures, predictors, potential confounders, and effect modifiers. Give diagnostic criteria, if applicable | | Not applicable to this study design |
| Data sources/ measurement | 8* | For each variable of interest, give sources of data and details of methods of assessment (measurement). Describe comparability of assessment methods if there is more than one group | | Not applicable to this study design |
| Bias | 9 | Describe any efforts to address potential sources of bias | 12<br>34-35 | Purposive sampling strategy: line 257-266<br>Strengths and limitations of study: lines 742-784 |
| Study size | 10 | Explain how the study size was arrived at | 11 | Line 237 |

Continued on next page

| | | | | |
|---|---|---|---|---|
| Quantitative variables | 11 | Explain how quantitative variables were handled in the analyses. If applicable, describe which groupings were chosen and why | 14 | Lines 301-302 'FGD and IDI participants provided verbal and written informed consent as well as their demographic data for descriptive analysis' |
| Statistical methods | 12 | (*a*) Describe all statistical methods, including those used to control for confounding | | Not applicable to this study design |
| | | (*b*) Describe any methods used to examine subgroups and interactions | | Not applicable to this study design |
| | | (*c*) Explain how missing data were addressed | | Not applicable to this study design |
| | | (*d*) *Cohort study*—If applicable, explain how loss to follow-up was addressed<br>*Case-control study*—If applicable, explain how matching of cases and controls was addressed<br>*Cross-sectional study*—If applicable, describe analytical methods taking account of sampling strategy | | |
| | | (*e*) Describe any sensitivity analyses | | Not applicable to this study design |
| **Results** | | | | |
| Participants | 13* | (a) Report numbers of individuals at each stage of study—eg numbers potentially eligible, examined for eligibility, confirmed eligible, included in the study, completing follow-up, and analysed | 15 | Line 305-314 (results section) |
| | | (b) Give reasons for non-participation at each stage | 15 | Line 309-310 |
| | | (c) Consider use of a flow diagram | | Not applicable to this study design |
| Descriptive data | 14* | (a) Give characteristics of study participants (eg demographic, clinical, social) and information on exposures and potential confounders | 15 | Line 305-312 (results section) |
| | | (b) Indicate number of participants with missing data for each variable of interest | | Not applicable to this study design |
| | | (c) *Cohort study*—Summarise follow-up time (eg, average and total amount) | | Not applicable to this study design |
| Outcome data | 15* | *Cohort study*—Report numbers of outcome events or summary measures over time | | Not applicable to this study design |
| | | *Case-control study*—Report numbers in each exposure category, or summary measures of exposure | | Not applicable to this study design |
| | | *Cross-sectional study*—Report numbers of outcome events or summary measures | | Not applicable to this study design |
| Main results | 16 | (*a*) Give unadjusted estimates and, if applicable, confounder-adjusted estimates and their precision (eg, 95% confidence interval). Make clear which confounders were adjusted for and why they were included | | Not applicable to this study design |
| | | (*b*) Report category boundaries when continuous variables were categorized | | Not applicable to this study design |

| | | |
|---|---|---|
| | (*c*) If relevant, consider translating estimates of relative risk into absolute risk for a meaningful time period | Not applicable to this study design |

Continued on next page

| | | | | |
|---|---|---|---|---|
| Other analyses | 17 | Report other analyses done—eg analyses of subgroups and interactions, and sensitivity analyses | | Not applicable to this study design |
| **Discussion** | | | | |
| Key results | 18 | Summarise key results with reference to study objectives | 30 | Lines 641-651 |
| Limitations | 19 | Discuss limitations of the study, taking into account sources of potential bias or imprecision. Discuss both direction and magnitude of any potential bias | 35 | Lines 773-784 |
| Interpretation | 20 | Give a cautious overall interpretation of results considering objectives, limitations, multiplicity of analyses, results from similar studies, and other relevant evidence | 35 | Lines 773-784 |
| Generalisability | 21 | Discuss the generalisability (external validity) of the study results | 34,35 | Lines 742-784 as appropriate to study design |
| **Other information** | | | | |
| Funding | 22 | Give the source of funding and the role of the funders for the present study and, if applicable, for the original study on which the present article is based | 36 | Included in funding statement, Line 810-815 |

*Give information separately for cases and controls in case-control studies and, if applicable, for exposed and unexposed groups in cohort and cross-sectional studies.
